# Neural Characteristics in Non-Suicidal Self-Injury and its Relation to Pain: A Functional and Structural MRI Study

**DOI:** 10.64898/2026.01.30.26345193

**Authors:** Karin B. Jensen, Sebastian Blomé, Jens Fust, Rosaleena Mohanty, Johan Bjureberg, Nitya Jayaram-Lindström, Eric Westman, Eva Kosek, Clara Hellner, William H. Thompson, Maria Lalouni

## Abstract

**Importance:** Pain is inherently aversive but provides emotional relief for individuals engaging in non-suicidal self-injury (NSSI). Despite the high prevalence and severity of NSSI, the neural mechanisms underlying pain processing in individuals with NSSI remain poorly understood.

**Objective:** To compare brain structure and functional connectivity between individuals with NSSI and controls and to relate brain function to pain inhibition.

**Design:** Cross-sectional, experimental.

**Setting:** MR Center at the Karolinska University Hospital in Stockholm, Sweden.

**Participants:** Women aged 18-35 years with NSSI (n=41) or matched healthy controls (n=40).

**Exposures:** Engagment in self-injury ≥ 5 days during the last year.

**Main outcomes and measures:** Magnetic Resonance Imaging (MRI) was used to examine brain structure and function related to pain regulation in individuals with NSSI (n=41) and healthy controls (n=40). The experimental pain test Conditioned Pain Modulation (CPM) was used to determine descending pain inhibition.

**Results:** We found higher connectivity between the brain’s somatomotor networks and subcortical areas during resting-state functional MRI in NSSI compared to controls (*P*=.009; Bonferroni corrected), particularly involving the thalamus and caudate nucleus. The connectivity was linked to the level of descending pain inhibition during CPM. There was no difference between NSSI and controls regarding brain morphometry.

**Conclusions and relevance:** Our findings suggest that individuals with NSSI may rely more on sensory-motor activations to regulate emotions. This study provides the first evidence linking specific brain circuits to pain regulation and self-injury behavior, highlighting potential pathways for more effective treatments for NSSI and related mental health conditions.

## Introduction

Pain is an aversive sensation that shapes our behavior and motivates us to avoid physical harm. However, some individuals intentionally inflict pain in order to reduce intense negative emotions, often referred to as non-suicidal self-injury (NSSI). For these individuals, pain serves a dual purpose as it acts both as an aversive experience and as a means of emotional relief.^1^ The prevalence of NSSI, like cutting and carving or burning the skin, is 17.7% among adolescents^2^ and 5.5% in adult community samples.^3^ While providing relief of negative emotions in the short-term, NSSI leads to suffering, disability, and potential escalation to serious harm or suicide attempts in the long-term.^4,5^ Despite the gravity and prevalence of self-injury, current treatments are not well-established as they lack empirical support,^6^ with limited evidence derived from a very small number of studies.^7^

To increase the understanding of mechanisms behind self-injury behavior, neuroimaging has been used to assess differences in brain structure and function between NSSI and healthy controls. Studies of NSSI participants with functional brain scans mainly included tasks focused on emotional regulation or reward processing,^8,9^ but some used painful stimuli.^10-12^ Few studies included participants with a current NSSI problem.

Previous studies have shown that patients with self-injury behavior have altered function in cortical and subcortical regions involved in emotional regulation and inhibititory cognitive processes^13, 14^. However, the studies has only allowed suggestive conclusions of functional differences between individuals with NSSI and healthy controls.. The neural mechanisms related to pain processing in individuals with NSSI, a core aspect of the behavior, remain largely unknown.

Experimental pain testing indicates that individuals with NSSI exhibit altered pain responses, including higher pain thresholds and pain tolerance, and a more effective decsending pain inhibition compared to healthy controls^15, 16.^ Furthermore, the risk of suicide is significantly higher in those who engage in self-harm^17^. Notably, individuals who experience little or no pain during NSSI are at twice the risk of suicide compared to those who feel substantial pain.^18^

Because altered pain processing is a hallmark of NSSI and linked to suicide risk, relating the neural circuitry to pain regulation in NSSI may provide critical insight into the biology underlying self-injury. Hence, the objective of this study was to link pain regulation to brain structure and function in women with NSSI compared to healthy controls using multimodal imaging. To this end, experimental pain testing was employed together with structural and functional magnetic resonance imaging (sMRI/fMRI). We assessed whether there were any differences between NSSI and controls in gray matter morphometry in the pre-specified regions of interest: pre-supplementary motor area, premotor cortex, intraparietal cortex, and temporoparietal junction, and assessed the functional connectivity between the somatomotor network and networks involved in salience signaling (subcortical and ventral attention).

## Methods and Materials

This study is part of a larger research project, investigating the role of pain regulation in women with NSSI. The first results from quantitative sensory testing and task-based fMRI have been reported elsewhere.^15^ The project was approved by the Regional Ethical Review Board in Stockholm (2018/1367-31/1). Data was collected between May 2019 and August 2020. The analyses were pre-registered on the Open Science Framework (OSF): https://osf.io/sbz4m, 2019; https://osf.io/72qfr, 2021. All study visits took place at the MRI Center at the Karolinska University Hospital in Stockholm, Sweden, between May 2019 and August 2020, with a disruption during the covid outbreak from March 2020 to May 2020. The study design was cross-sectional, comparing questionnaire data, experimental pain tests, and neuroimaging (functional and structural) between NSSI participants and a matched group of contols.

### Participants

Based on Koenig et al.,^16^ 29 participants per group were needed for 80% power to detect pain threshold differences which has been presented elsewhere^15^. To allow for smaller effects in other measures, we enrolled 41 NSSI participants and 40 controls. Participants eligible for inclusion were right-handed women aged 18-35 years, either engaging in NSSI, or healthy controls. The groups were matched regarding age and educational level. The upper age limit was chosen because NSSI is more prevalent in younger ages and to ensure a homogenous sample. NSSI participants were recruited via flyers posted in waiting rooms at outpatient psychiatric clinics and via social media advertisements. Controls were recruited via social media advertisements (Facebook). Inclusion criteria for NSSI participants specifically: a) self-injury ≥ 5 days during the last year. Exclusion criteria for both groups: b) chronic inflammatory, autoimmune, or other somatic disorders requiring treatment, c) pain condition, d) contraindication for fMRI (e.g., metal implant, pregnancy), e) suicide attempts during the last year, f) suicidal plans or acute risk for suicide. Exclusion criteria for controls specifically: g) self-injury behavior during the last year, and h) ongoing treatment for depression or anxiety. Individuals were interviewed via telephone to assess their eligibility for the study. NSSI participants met once with a licensed psychologist to assess study risk and psychiatric comorbidity, then completed pain tests across two consecutive visits alongside controls.

### Outcome measures

#### Conditioned Pain Modulation

Conditioned pain modulation (CPM) is a validated experimental method for measuring descending pain inhibition in a non-invasive way. For two summaries of brain mechanisms linked to CPM see two reviews.^19,20^ In the present study, pressure pain thresholds (kPa) were measured on the left calf (location marked by a pen) while ischemic pain was induced on the right forearm, with participants sitting upright. Pressure pain theresholds were defined as the pressure applied when an increasing pressure was experienced as slightly painful. Pressure stimuli were applied via a hand-held algometer (1 cm^2^ probe), and ischemic pain was induced using a blood pressure cuff (Hokanson Inc, Bellevue, WA, USA) and a 1 kg dumbbell, which participants flexed up and down (1 movement per second). Ischemic pain was applied until participants indicated that their forearm pain was 7 on a 0-10 scale (0=no pain, 10=worst imaginable pain). Two pressure pain thresholds were applied before the ischemic pain was induced and two pressure pain thresholds were induced simultaneous with the ischemic pain at level 7. The participants were asked to focus specifically on the pressure pain thresholds while they were appliedThe reduction in pain via CPM was calculated as the mean difference in pressure pain thresholds (kPa) on the calf with versus without concurrent ischemic pain. Data from pain testing was available from all participants (NSSI n=41; controls n=40).

#### Questionnaires and NSSI characteristics

Questionnaires were self-assessed by all participants in an online data collection platform (BASS, eHealth Core Facility, Karolinska Institutet, Sweden). Questionnaires about borderline symptoms (short version of the Borderline Symptom List; BSL-23),^21^ self-criticism (the Self-Rating Scale; SRS),^22^ and difficulties with emotion regulation (brief version of the Difficulties with Emotion Regulation Scale; DERS-16)^23^ were self-assessed. Frequency and function of NSSI were assessed with the Functional Assessment of Self-Mutilation (FASM)^24^ survey. Frequency was indicated by number of days with self-harm behavior during the last year with the alternatives: 1-4, 5-15, 15-50, 50-up. NSSI duration was assessed by subtracting the participants’ debut age of NSSI from their age.

#### Magnetic resonance imaging (MRI)

A 3T General Electric 750 MRI scanner was used to acquire a high-resolution BRAVO 3D T1-weighted image sequence (1 × 1 × 1-mm voxel size, 176 slices) of the brain. Functional imaging data during resting-state (participants lying still in the scanner with eyes open) were acquired using a gradient echo-planar imaging sequence (EPI), sensitive to blood-oxygenation-level-dependent (BOLD) contrast; repetition time = 2.2 ms, echo time = 30 ms, field of view = 230 x 230mm, flip angle = 80°, slice thickness = 3 mm. Images included coverage of the whole-brain. Eight minutes of resting-state EPI images were collected for each participant, resulting in 220 brain volumes for use in analyses of functional brain connectivity.

Resting-state imaging was available for n=33 participants with NSSI and n=40 controls. Structural MRI data was available for n=37 of participants with NSSI and n=39 controls. The missing participants were due to: Not being able to remove metal object from body (n=1), claustrophobia (n=1), declined MR (n=1), removed from sMRI analysis because of insufficient quality of images (n=2), and missing resting-state fMRI data because of technical difficulties (n=5).

#### Functional brain analyses

The functional brain images were quality assessed using MRIQC.^25^ Framewise displacement was used to determine if there were any excess movement during image acquisition. No participants were excluded from the analysis as there were no framewise displacement (FD) over >0.5 in >15% of the images. The data was preprocessed using fMRIPrep 20.2.0.^24^ It provides minimal preprocessing, including motion correction, field unwarping, normalization, bias field correction, and brain extraction. Lastly, preprocessed images were denoised using the 36-parameter strategy during the parcellation step using nilearn.^27^

To determine functional brain networks, the Schaefer parcellation atlas was used^28^ in which 400 cortical nodes are mapped to 7 different brain networks.^29^ The Oxford-Harvard subcortical atlas was used to determine a subcortical brain network (15 nodes), normalized to MNI152NLin2009cAsym (https://identifiers.org/neurovault.collection:262). This resulted in a total of 415 nodes categorized into 8 defined resting-state networks, 7 cortical and one subcortical. The cerebellum was not included in our analyses because of inadequate coverage during image acquisition.

The parcellated brain data were pairwise correlated using Pearson correlation, and aggregated into the eight networks (visual, somatomotor, dorsal attention, ventral attention, limbic, frontoparietal control, default mode, and the subcortical network). A priori defined analyses of the connectivity between the somatomotor network and those networks implicated in salience signaling (subcortical and ventral attention) were performed. The link between CPM and brain connectivity was analyzed using Pearson correlations.

Independent samples t-test was used to determine whether there was a significant difference in network connectivity between groups (NSSI versus controls). This analysis was corrected for multiple comparisons (all network pairs) using Bonferroni correction.

#### Structural brain analyses

The T1-weighted images were preprocessed in Freesurfer version 7.1.1 through the HiveDB database system.^30^ Quality control of the T1-weighted images was performed based on coverage and white and gray matter differentiation. Standard atlases proposed by Desikan^31^ and Fischl^32^ were used to correspond the regions of interest with the Freesurfer output. The predefined Freesurfer cortical areas included in the analyses were precentral, caudal middle frontal, postcentral, superior parietal, inferior parietal, and banks of the superior temporal sulcus. Right and left hemisphere were analyzed separately. To correct for differences in head sizes, normalized values for volumes were used. These were obtained by dividing the values for volume by the estimated total intracranial volume (eTIV). Generalized linear models were used to assess differences in gray matter volume and cortical thickness between the NSSI and the control group (whole brain analyses) and false discovery rate was used to control for multiple comparisons. T-tests were used to assess differences in volume and cortical thickness in the regions of interest between groups.

## Results

### Participants

Out of 107 individuals with NSSI and 104 controls assessed for eligability, 41 NSSI participants and 40 controls were included. A total of 108 individuals were excluded, n=52 in the NSSI group (suicidality n=8, pain condition n=16, not fulfilling NSSI criteria n=21, contraindication for MR n=7), and n=56 in the control group (ongoing NSSI n=16, pain condition n=7, contraindication for MR n= 22, psychiatric treatment n=11). Also, n=14 NSSI individuals and n=8 controls declined to participate. Mean age was 23.4 (SD=3.9) years. For detailed demographic and clinical characteristics, see **Table 1**. Within the NSSI group, the most common self-injury behaviors were hitting (n=32), cutting (n=31), or carving (n=31), with a mean number of methods of 5.6 (SD = 2.2), range 2–12. The most reported reasons for self-harm were “to punish oneself” and “to stop bad feelings”. The frequency of NSSI behavior during the last year was 50 days or more reported by 30%, 15-50 days reported by 45%, and 5-15 days reported by 25% of the participants with NSSI. The mean age at onset of NSSI was 13.2 (SD=3.1) years and the average duration of NSSI was 10 years. Psychiatric comorbidity was reported by 32 out of 39 participants with NSSI. The most common diagnoses were depression (n=21), anxiety disorder (n=12), and borderline personality disorder (n=11).

**Table 1.** Demographic and clinical characteristics.

|  | NSSI | Controls | Total | Statistic | P-value |
| --- | --- | --- | --- | --- | --- |
| **Number of participants** | 41 | 40 | 81 |  |  |
| **Age, mean (SD), years** | 22.7 (3.6) | 24.0 (4.1) | 23.4 (3.9) | t = -1.54 | .128 |
| **Educational level, n (%)** |  |  |  |  |  |
| **Elementary school** | 7 (17.1) | 7 (17.5) | 14 (17.3) | $\chi^2 = 1.19$ | .755 |
| **High school** | 27 (65.9) | 24 (60.0) | 51 (63.0) |  |  |
| **College degree** | 5 (12.2) | 8 (20.0) | 13 (16.1) |  |  |
| **Other** | 2 (4.9) | 1 (2.5) | 3 (3.7) |  |  |
| **Clinical measures** |  |  |  |  |  |
| **BSL-23, mean (SD)** | 2.3 (0.9) | 0.4 (0.3) | 1.3 (1.2) | t = -13.0 | <.001 |
| **SRS, mean (SD)** | 33.7 (8.5) | 16.2 (7.9) | 25.0 (12.0) | t = -9.7 | <.001 |
| **DERS-16, mean (SD)** | 56.2 (13.3) | 27.1 (9.9) | 41.8 (18.8) | t = -11.2 | <.001 |
| Abbreviations: SD, standard deviation; NSSI, nonsuicidal self-injury; BSL-23, Borderline Symptom List; SRS, Self-Rating Scale (self-criticism); DERS-16, 16 Item Version of the Difficulties in Emotion Regulation Scale. |  |  |  |  |  |

### Brain function

#### Resting-state functional magnetic resonance imaging

When comparing the NSSI group with controls, there was a significant difference in resting-state functional connectivity between the somatomotor (Figure 1A) and subcortical networks (Figure 1D), with NSSI participants displaying a stronger functional connectivity compared with controls; t = −3.40; *P*=.009; Bonferroni corrected (Figure 1B-C). No other comparisons survived the significance level when adjusting for multiple comparisons. To unpack which anatomical structures within the subcortical network that contributed to the significant difference, exploratory analyses where each subcortical region was treated as a region of interest were displayed as density plots (Figure 1E). The density plots indicate that the thalamus and the caudate nucleus specifically contributed to the difference in connectivity between the somatomotor and subcortical networks.

**Figure 1.**
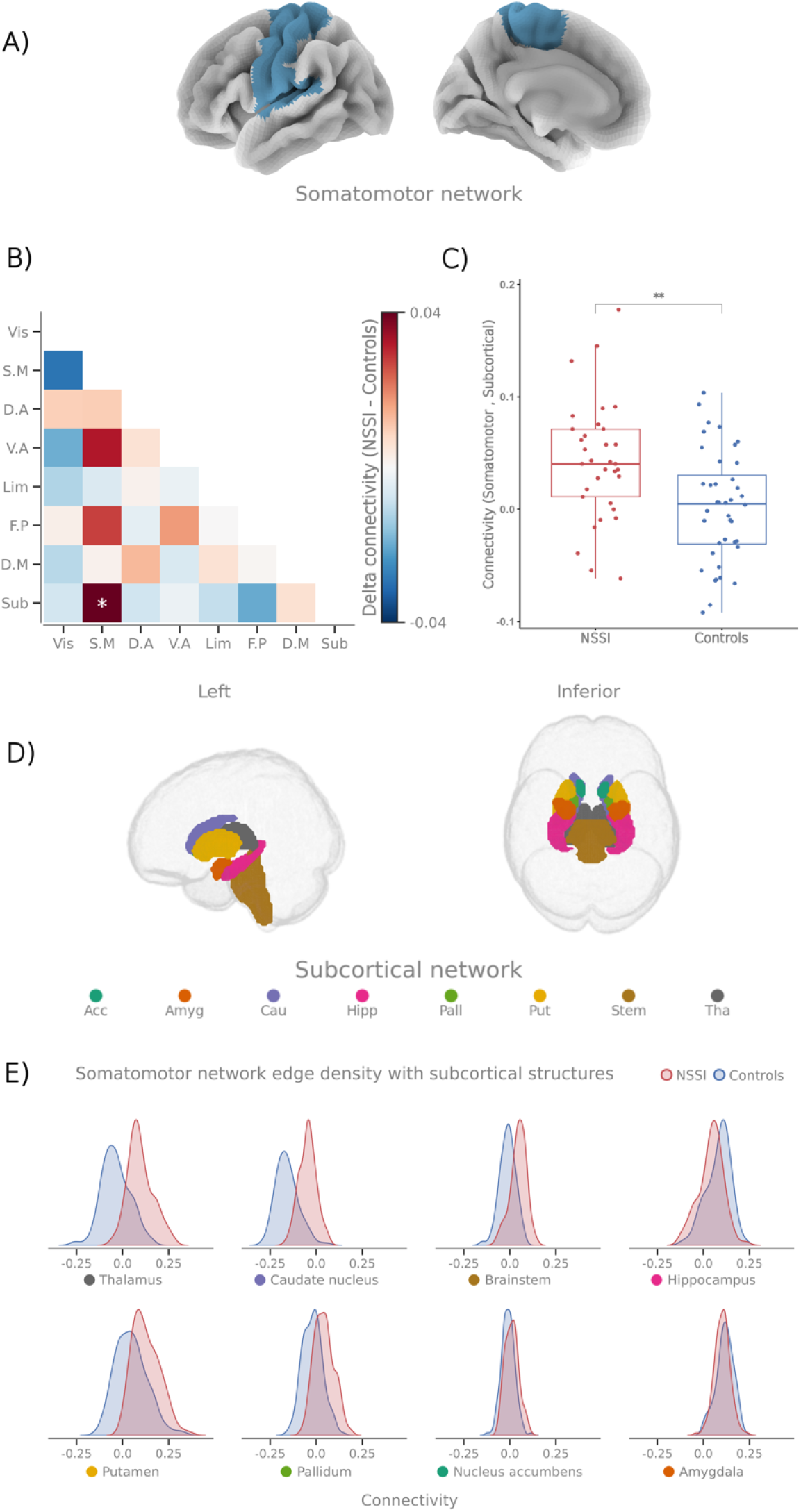
Brain connectivity results. **A**. Graphic representation of the somatomotor network. **B**. Connectivity matrix representing the difference in functional connectivity between the networks (NSSI - Controls) NSSI=Non-suicidal self-injury, Vis=Visual, S.M=Somatomotor, D.A=Dorsal attention, V.A=Ventral attention, Lim=Limbic, F.P=Frontoparietal control, D.M Default mode, Sub=Subcortical. **C**. Barplot highlighting the difference in subcortical- and somatomotor network connectivity between groups (*P* =.001). **D**. Graphic representation of the brain regions included in the subcortical network (thalamus, caudate nucleus, brainstem, hippocampus, putamen, pallidum, nucleus accumbens, amygdala). **E**. Density plots for the somatomotor functional connectivity with each anatomical structure in the subcortical network.

#### Resting-state fMRI functional connectivity and conditioned pain modulation

NSSI participants displayed greater inhibition of pain than controls, as indicated by larger degree of conditioned pain modulation during experimental testing (196 versus 102 kPa, respectively; *P*<.001). Across groups, there was a significant association between the somatomotor-subcortical functional connectivity and conditioned pain modulation (t = 2.09, df = 68, *P*=.040), but no significant interaction between groups.

### Brain structure

Cortical thickness analyses revealed one difference in brain structure between NSSI participants and controls, located in the caudal middle frontal gyrus (right hemisphere), representing a portion of the premotor cortex. The cortical thickness was thinner in NSSI participants (2.54 mm) than in controls (2.62 mm), *P*=.042. This finding did, however, not survive correction for multiple comparisons. No other measures of brain structure passed an uncorrected threshold for statistical significance.

## Discussion

In the present study comparing brain structure and functional connectivity between individuals with NSSI and controls, we provide evidence for increased resting-state functional connectivity between the somatomotor and subcortical brain networks in individuals with NSSI compared to controls. These results led us to further explore the specific subcortical areas involved. Our exploratory analyses indicate that the thalamus and caudate nucleus accounted for the greatest connectivity difference among the subcortical structures. Unlike other studies that included participants with borderline personality disorder or a history of self-injury^33^ this study focused on individuals with current NSSI. Based on the pronounced self-injury behaviors in the present group, including severe physical harm, we hypothesized that neural alterations would be reflected in the somatosensory and motor regions of the brain. In line with our hypothesis and preregistered analyses, we found differences in the functional organization of the brain’s somatomotor networks and subcortical areas, but no evidence for differences in brain structure. Additionally, the connectivity between the somatomotor areas and subcortical regions was linked to the level of endogenous pain inhibition. To our knowledge, these findings provide the first evidence of specific brain circuitry linking pain regulation to self-injury. Understanding this connection may help identify risk factors and improve prevention by incorporating pain profiles into psychiatric assessments.

### Emotional regulation versus pain regulation

The primary reason for NSSI is difficulty in regulating negative emotions,^34, 35^ and in the present study participants with NSSI reported greater difficulties with emotional regulation compared to controls.^15^ Despite difficulties in regulating negative emotions, our findings indicate that individuals with NSSI exhibit enhanced pain regulation, as shown by augmented inhibition of pain during conditioned pain modulation.^15^ This suggests that NSSI is not linked to a general lack of inhibitory control. Instead, our data suggest increased inhibition of pain-specific signals, reflected in increased somatomotor-subcortical connectivity. While emotion regulation typically involves prefrontal-limbic communication,^36^ individuals with NSSI may use a previously unexplored somatomotor-subcortical circuitry to down-regulate both pain and emotion, triggered by noxious stimulation to their bodily tissues.

### The specific subcortical regions in NSSI brain circuitry

The enhanced somatomotor-subcortical connectivity observed in NSSI participants was most prominent in the thalamus and caudate nucleus. The thalamus is central for filtering afferent sensory input,^37^ and the stronger thalamus-somatomotor connectivity may reflect augmented sensory processing in NSSI participants, e.g., that somatosensory information is more salient in NSSI. Enhanced functional connectivity between the thalamus and the somatomotor network has also been reported in different psychiatric disorders^38, 39^ and could thus be a more general marker of psychiatric ill-health, not specific for NSSI. The caudate nucleus, part of the dorsal striatum, is considered a central hub in circuits involved in decision-making and linking emotional and motivational states to motoric action.^40^ Abnormalities in caudate nucleus function has previously been detected across a variety of psychiatric disorders in which NSSI is prevalent,^41^ e.g., bipolar disorder and major depressive disorder, making it a key focus in understanding NSSI. Other findings from brain imaging studies during reward-related tasks suggest altered activation of the caudate nucleus in NSSI,^42, 43^ interpreted as altered function of reward anticipation. The present study, with a comparably large number of participants, found evidence for stronger connectivity between the somatomotor network and the caudate nucleus in participants with NSSI. We suggest that this may reflect an increased reliance on sensory processing, and potentially sensory attenuation,^44^ to regulate emotions, in addition to prefrontal control circuitry.

The significant correlation between CPM and the strength of somatomotor-subcortical connectivity is highly interesting even if there was no apparent difference between NSSI and healthy controls. The connectivity between the somatomotor and subcortical brain structures may still be valuable for future exploration of the role of sensory processing in emotional regulation. Thus, our results should be validated in future studies where CPM is performed by blinded experimenters and the addition of a formal control for attention and distraction during testing, as suggested by Kennedy et al. (2016).^45^

### Brain Structure

A previous study found differences in gray matter morphometry between individuals with NSSI and controls in the insula and the anterior cingulate cortex.^46^ In patients with borderline personality disorder, in which NSSI is common, several studies have found structural brain differences in the cortical and limbic regions between patients and controls.^47^ However, a large study using two community samples found no structural differences related to level of borderline personality traits,^48^ and a recent meta-analysis failed to find consistent findings.^14^ In the present study we only found one structural difference in the right caudal middle frontal gyrus, which did not survive correction for multiple comparisons.

### Limitations

In the control group, participants with ongoing treatment for psychiatric disorders were excluded. At the same time, most participants with NSSI reported that they had one or more psychiatric disorder. Thus, it cannot be completely ruled out that the results found in our study are confounded by the presence of a psychiatric condition, even if a recent study among individuals with major depressive disorder, some of which also presented with NSSI, suggested specific brain alterations linked to NSSI.^49^ Other limitations are the cross-sectional study design which precludes causal inferences.

## Conclusion

Our results demonstrate a more enhanced somatomotor-subcortical functional connectivity in NSSI participants compared with controls, which might suggest a reliance on sensory processing to regulate pain and emotions, reflected in the thalamus and caudate nucleus.

## Data Availability

The datasets presented in this article are not readily available because we do not have ethical permit to share the data. Requests to access the datasets should be directed to.

## Acknowledgements

We thank all the participants of the study, the psychiatric units that helped us inform about the study, and the patient organization SHEDO for their support.

